# Connectome-based predictive modeling of early and chronic psychosis symptoms

**DOI:** 10.1101/2024.05.20.24307572

**Authors:** Maya L. Foster, Jean Ye, Albert Powers, Nicha Dvornek, Dustin Scheinost

**Affiliations:** Department of Biomedical Engineering, Yale University; Interdepartmental Neuroscience Program, Yale School of Medicine; Department of Psychiatry, Yale School of Medicine; Wu Tsai Institute, Yale University; Department of Radiology & Biomedical Imaging, Yale School of Medicine; Department of Statistics & Data Science, Yale University; Child Study Center, Yale School of Medicine

## Abstract

The symptoms of psychosis-spectrum disorders, which include positive symptoms (e.g., hallucinations and delusions) and negative symptoms (e.g., memory impairment and disorganized thinking), can cause significant distress and disability. Despite shared symptomatology and converging brain correlates, early (EP) and chronic (CP) psychosis differ in their symptom-related treatment response. At present, the mechanism underlying these differences is unknown, in large part because EP and CP have predominantly been studied and characterized independently or in comparison to control populations. To answer this question, we use connectome-based predictive modeling (CPM) and resting-state functional magnetic resonance imaging to identify biologically-based early (EP, n=107) and chronic (CP, n=123) psychosis symptom networks. We predicted both samples’ total, positive, and negative symptoms from the PANSS. Virtual lesioning analyses highlight the frontoparietal network as a critical component of EP and CP symptom networks, but the specific functional connections used for prediction differ. Finally, group differences compared to healthy controls (n=150) were observed for CP but not EP. These differences broadly overlapped with the symptom model for both EP and CP. Our results encourage using longitudinal studies to track connectivity changes in putative symptom networks during the progression of psychosis, as they may be explicative of EP-CP treatment differences.

## Introduction

Schizophrenia and other psychosis-spectrum disorders affect 1% of the world’s population (Felix 2020). Patients face both positive (e.g., hallucinations and delusions) and negative (e.g., blunted affect, avolition, and anhedonia) symptoms that cause significant distress and disability (Malla and Payne, 2005; Griffiths et al., 2019; Hopeway, 2023; American Psychiatric Pub, 2013).

Negative symptoms often arise first in the prodromal period before an official clinical diagnosis (Larson et al., 2010; Strauss et al., 2013; Correll and Scholler 2020; Calabrese and Khalili, 2024). From a clinical standpoint, the evolution of positive symptoms into fully formed and disruptive symptoms indicates conversion to early psychosis (EP) (McGorry et al., 2008; Larson et al., 2010). From then on, symptom courses are patient-dependent and consist of remission, sustained, improved, or progressive worsening (Heilbronner et al., 2016; Correll and Scholler 2020; Starzer et al. 2023). The extent of symptom burden or relapse occurrence (Bottai et al., 2009) over time is what distinguishes EP from chronic psychosis (CP).

Identifying the neural correlates of psychosis symptoms is critical because the dominant treatment and diagnosis strategy for this condition is symptom-centric (Taminga, 2001; Calabrese and Khalili, 2024). Additionally, identifying symptoms-specific brain networks would be an asset to enhancing and optimizing care and treatment (Zhu et al., 2023). Psychosis symptoms are thought to be a consequence of “disruptions of the coordinated functioning of distributed brain regions” (Fu et al., 2021). They potentially result from broad alterations of the default mode network (DMN) (Garrity et al., 2007, Karlsgodt et al., 2010, Dong et al., 2018, Fu et al. 2021), subcortical (Dong et al., 2018, Fu et al. 2021), cerebellar (Yue et al, 2016, Fu et al. 2021), frontoparietal (Dong et al., 2018), thalamocortical (Anticevic et al., 2014, Dong et al., 2018), salience (Dong et al., 2018), somatosensory (Dong et al., 2018), and sensorimotor networks (Fu et al. 2021). Abnormal dynamic functional connectivity (Cattarinussi et al., 2023) and salience-default mode functional connectivity (Hare et al., 2019) are associated with positive and negative symptoms in schizophrenia. Decreased cerebellar activation during a verbal Stroop task (Vanes et al., 2019) and altered resting-state cerebellar-cerebral functional connectivity (Choi et al., 2023) have been associated with negative symptom burden in EP patients. Aberrant functional connectivity in the frontotemporal (Abram et al., 2016), cerebellar-prefrontal (Brady, Jr et al., 2019), and frontostriatal (Shukla et al., 2019) areas are observed in CP.

Despite shared symptomatology and converging structural (Pantelis et al., 2005, Luvsannyam et al., 2022) and functional (Vanes et al., 2019) brain correlates, EP is substantially more responsive to symptom treatment than CP. Indeed, early intervention (within five years of disease onset) has proven to enable better treatment outcomes (van Schalkwyk et al., 2015) for psychosis. The reason for these differences remains open, which could stem from a range of stage-delineating factors such as the presence of physical comorbid disorders (Zierotin et al., 2023), antipsychotic medication exposure (Lewandowski et al. 2020), and symptom burden (Vanes et al., 2019). Previous studies have identified brain-based biomarkers that could explain treatment differences, such as increased connectivity between the salience and the default mode networks and between the salience network and the cerebellum in EP (Mallikarjun et al., 2018) and reduced frontotemporal (John H. John 2009), frontothalamic (John H. John 2009, Hwang et al. 2021), and thalamocortical connectivity (Anticevic et al., 2014, Welsh et al, 2010) in CP. Nonetheless, these are not sufficiently explanatory. Further isolating the neurobiological correlates associated with symptom burden and identifying the neural bases of EP-CP delineating factors at a whole brain level could help researchers understand the mechanisms supporting early intervention success in EP, inform preventative treatment development, and stage-based therapeutic strategies (McGorry et al., 2008).

Predictive models are well-suited for this task, as they can provide insights into symptoms-associated connections at the individual level and can reduce overfitting (Scheinost et al., 2019). Connectome-based predictive modeling (CPM) is a well-established, data-driven method for predicting symptoms from connectomes or whole-brain functional connectivity data (Shen et al., 2017). CPM has successfully predicted several phenotypes across psychiatric conditions (Yip et al., 2019, Li et al., 2023, Yoo et al., 2018, Tejavibulya et al., 2022). Prior success primes CPM as a promising tool for investigating symptom networks specific to or overlapping between EP and CP.

In this study, we identify biologically-based EP (Lewandowski et al., 2020) and CP (Tanaka et al., 2021) psychosis symptom networks using CPM and resting-state functional magnetic resonance imaging (rsfMRI). We hypothesized that EP and CP impact similar system-level networks with distinct edge topography due to phenotypic overlap but severity differences. Additionally, we hypothesized that networks predicting symptoms would include the DMN (Bastos-Leite et al., 2015; Wang et al., 2015; Hu et al., 2016; Hilland et al., 2022; Chen et al., 2023; Fan et al., 2022) and frontoparietal network (FPN) (Briend et al. 2020).

## Methods

### Participants

We used two publicly available resting-state fMRI datasets (see Table 1 for participant demographics), the Human Connectome Project Early Psychosis (HCP-EP) (Lewandowski et al. 2020) and the Strategic Research Program for Brain Sciences (SRPBS) Multi-disorder Connectivity Dataset (Tanaka et al., 2021). The HCP-EP dataset consists of early psychosis (EP) patients (within the first five years of the first presentation of psychosis symptoms) and healthy controls (HCs) from four different recruitment sites. The HCP-EP sample consisted of affective (AP) and non-affective psychosis (NAP) patients, who were diagnosed according to the DSM-V (American Psychological Association, 2013). The SRPBS dataset is a transdiagnostic population of patients with psychosis symptoms of varying severities. We included HCs and individuals diagnosed with schizophrenia spectrum disorder (SSD) from three different recruitment sites in Japan (Kyoto University, Showa University, and University of Tokyo Hospital). Participants in both datasets underwent resting-state functional magnetic resonance imaging (rsfMRI) and measured symptom severity with the Positive and Negative Syndrome Scale (PANSS) (Kay et al. 1987, Figure S1).

### Connectome-based predictive modeling (CPM)

After standard resting-state preprocessing (see Supplementary Methods), we built CPM models to predict psychosis symptom scores using resting-state connectomes for EP and CP separately (Shen et al., 2017). During quality control for the SRPBS dataset, 26 brain nodes were excluded due to incomplete slice coverage (Figure S2). Our primary outcome is the PANSS total score because it combines information across all symptom score dimensions. Secondary results include predicting the negative and positive symptoms and the general psychopathology score (GPS). Feature selection included one hundred iterations of 10-fold cross-validation with a significance threshold of p<0.05 and partial correlation for controlling covariate confounds (Hsu et al., 2018). EP and CP models co-varied for site, sex, age, and motion. EP models also co-varied for medication exposure. As the HCP-EP had individual PANSS items, we investigated if a particular item was driving symptom prediction (see Supplement Methods).

Prediction performance was evaluated using Pearson’s correlation (r) and mean square error (MSE). We report the median Pearson’s correlation across the one-hundred 10-cross-validated models. Significance was determined with permutation testing. A null distribution of Pearson’s r values was produced by randomly shuffling the link between an individual’s symptom and connectivity matrix. Nonparametric p-values were calculated as follows: (p=(#[r_null>r_median]+1)/1000). Here, #[r_null>r_median] indicates the number of instances of permuted predictions had a larger Pearson’s correlation than the median of the non-permuted predictions. Because only a positive association between predicted and actual values indicates prediction above chance (negative associations indicate a failure to predict), one-tailed p-values are reported.

### Anatomical characterization of models

We characterized the anatomy of the symptom models with three approaches. Each CPM model consists of a positive network (i.e., a set of edges where increased connectivity is positively associated with symptom severity) and a negative network (i.e., a set of edges where increased connectivity is negatively associated with symptom severity). The positive and negative networks are characterized independently. First, we characterized the distribution of intrahemispheric and interhemispheric edges. Second, we classified edges based on the distance between the nodes (short, mid-short, mid-long, and long-distance edges) and quantified these for each model. Third, we conducted virtual lesion analyses to assess which functional networks are important for prediction (see Supplement Methods).

### Comparisons between EP and CP models

We compared EP and CP models using two approaches. First, we generalized the median-performing models across cohorts (i.e., applying a model trained in EP to test in CP and vice-versa). The model applied for cross-prediction was a consensus model that consists of edges that are a member of each of the ten cross-validated runs for the median performing model for each symptom. Generalizability was assessed using Pearson’s correlation. Significant generalization suggests that models are incorporating similar neurobiological information. Second, we determined the similarity of model features at the edge, node, and network levels. Significant for edge-level was based on the hypergeometric cumulative distribution 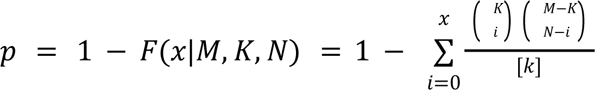, where *x* equals the number of overlapping edges between the EP and CP models, *K* equals the number of edges in the EP model, *n* equals the number of edges in the CP model, and *M* equals the total number of edges in the matrix (35,778). Pearson’s correlation was used to compare the node and network-level contributions between models.

### Group differences

We investigated group differences in connectomes between EP or CP and HCs. The network-based statistic (NBS) was used to correct for multiple comparisons (Zalesky et al., 2010). Similar to the above, we investigated how these group differences overlapped with the symptom models at the node and network levels (see Supplementary Methods).

## Results

### Participant characteristics

Participants were included for primary analysis if they had complete, non-zero scores and rsfMRI data with an average head motion <0.2 mm. Quality-controlled participants (N=383) included n=150 HCs (HCP-EP=57, SRPBS=93), n=107 EP, and n=123 CP. PANSS scores were significantly higher in CP than in EP (Table 1). Race and site information is shown in Tables S1-2.

### Predictive modeling of psychosis symptoms in EP

CPM successfully predicted total scores (r=0.31, MSE=338.95, p<0.001, 1-tailed permutation testing, 100 iterations, Figure 1), positive scores (r=0.29, MSE=26.14, p<0.001, 1-tailed permutation testing, 100 iterations), and negative scores (r=0.29, MSE=37.93, p=0.02, 1-tailed permutation testing, 100 iterations). Controlling for age, sex, head motion, medication exposure, phenotype, and site did not impact prediction (see Table S3). The GPS model was insignificant (Table S3). The suspiciousness and grandiosity items drive the positive symptom model prediction. In contrast, removing the hallucination and delusion items improved prediction (Table S4).

### Anatomical characterization of symptom models in EP

Here, we focus on the total score model. Similar characterizations for positive and negative symptom models are found in the supplement (Table S5-6). The positive network consisted of 312 edges. The negative network consisted of 349 edges (see Figure 2). Predictive edges span the whole brain and every canonical network. No hemispheric differences existed for the positive or negative networks (Table S5). No differences existed in the distribution of long-range and short-range connections for either the positive or negative networks (Table S6). Only lesioning the FPN worsened the total score prediction (Table S7), suggesting the FPN was the most responsible for total score prediction.

### Predictive modeling of psychosis symptoms in CP

CPM successfully predicted total (r=0.31, p<0.001, MSE=338.95, 1-tailed permutation testing, 100 iterations), positive (r=0.29, p<0.001, MSE=26.14, 1-tailed permutation testing, 100 iterations), negative (r=0.29, p=0.02, MSE=37.93, 1-tailed permutation testing, 100 iterations), and general psychopathology scores (r=0.27 p<0.001, MSE=89.10, 1-tailed permutation testing, 100 iterations; Figure 3). Controlling for age, sex, head motion, medication exposure, phenotype, and site did not impact prediction (Table S4).

### Anatomical characterization of symptom models in CP

As above, we focus on the total score model. Similar characterizations for positive, negative, and general psychopathology symptom models are found in the supplement (Table S8-9). The positive network consisted of 379 edges. The negative network consisted of 342 edges (Figure 4). Predictive edges span the whole brain and every canonical network (refer to Supplement). The positive network had significantly more edges in the right hemisphere compared to the left (right: 103, left: 51, χ2=5, p=0.03) and between hemisphere compared to within (between: 225, within: 154, χ2=6.35, p=0.01). No hemispheric differences existed for the negative network (Table S5). No differences existed in the distribution of long-range and short-range connections for either the positive or negative networks (Table S6). Virtual lesioning did not worsen the total score prediction (Table S9), suggesting that all functional networks contribute equally to total score prediction.

### Similarity of EP and CP symptom models

The EP positive symptom model predicted CP positive symptoms (r=0.23, p=0.01, df=121, see Table S10). All other symptom models failed to predict between cohorts (Table S10-11). At the edge level, there was no significant overlap between the EP and CP symptom models (p’s>0, r’s=-0.0032-0.0171, Table S12). At the node level, all EP and CP symptom models were correlated (r’s>0.13), except for the positive network of the positive symptom model (r=0.08, Table S13). At the network level, EP and CP symptom models were correlated (r’s>0.58, Table S14).

### NBS-derived group differences between patients and healthy controls

NBS revealed that at the group level, CP—but not EP—significantly differed from matched HCs (p<0.05, corrected, Table S15). Increased connectivity for CP was located in the FPN, somatomotor, and salience networks. Decreased connectivity for CP was located in the FPN, somatomotor, and salience networks. These case-control differences broadly overlapped with the symptom model for both EP (r’s>0.48, Table S16) and CP (r’s>0.58, Table S17).

## Discussion

We used CPM to identify biologically-based networks that underpin positive and negative symptoms in early (EP) and chronic psychosis (CP). PANSS positive and negative were predicted in EP; PANSS positive, negative, general psychopathology, and total symptom scores were predicted in CP. Models were robust to covariates (e.g., sex, age, medication exposure). While these models were complex—with contributions from every canonical brain network, the frontoparietal network (FPN) emerged as a critical predictor of symptoms across analyses. Models were similar between EP and CP at the system level (i.e., node and network). Nevertheless, the specific functional connections differ across groups. Finally, group differences compared to healthy controls (n=57) were observed for CP but not EP.

Psychosis and its cardinal symptoms likely emerge from whole-brain rather than focal disruptions. Accordingly, the literature reports a wide range of regions and networks altered in psychosis. Compared to controls, literature reports altered connectivity in the frontal lobes, the parietal cortex, the cerebellum, and the anterior cingulate gyrus (Spence et al., 2000; Whalley et al., 2005; Whitfield-Gabrieli et al., 2009, Woodward et al., 2009, Li et al., 2010) in high genetic-risk for developing schizophrenia populations. FPN (E. Fuller Torrey 2007, Woodward et al., 2009), motor network (Bernard et al., 2014; Bernard et al. 2017), insular cortex (Palaniyappan and Liddle, 2012), and medial prefrontal cortex (Pomarol-Clotet et al., 2010) dysfunction are also implicated in schizophrenia etiology and psychosis symptomatology. Our results echo these findings, as every brain region and network contributed to our models.

Still, only lesioning the FPN significantly impacted the prediction, highlighting its importance, particularly in negative symptoms. The disruption and dysfunction of the FPN are well-documented as hallmarks of psychosis (Yildiz et al., 2007; E. Fuller Torrey, 2007; Woodward et al., 2009; Yildiz et al., 2011; Roiser al., 2013, Briend et al., 2020). The FPN is a component of the cognitive control system and a flexible hub (Cole et al., 2014). It is pivotal in directing real-time task demands (Cole et al., 2014) and goal-directed behavior (Marek and Dosenbach 2018). Importantly, many negative symptoms are rooted in the hubs of the FPN, such as the dorsolateral prefrontal cortex and the anterior inferior parietal lobule. Our findings align closely with this existing literature.

Across analyses, results were similar between EP and CP at the system (i.e., node and network), not edge level. Further, we could predict positive symptoms in CP from the EP model. In other words, the functional anatomy underlying EP and CP is similar when looking at the forest. However, there are differences when looking at the implicated trees. These results are likely due to phenotypic overlap but differences in severity and disease progression. For example, they are broadly consistent with the neuroprogression theory of psychosis (Lewandowski et al., 2020), where significant cumulative brain changes occur during the years following an initial episode. Alternatively, this result may indicate that individual edges in a network are correlated and contain redundant information (Faskowitz et al, 2020; Jo et al., 2021; Novelli et al., 2022; Rodriguez et al., 2022). That one edge in a network is included in a model versus another may be due to data idiosyncrasies unrelated to psychosis neurobiology.

Also, across analyses, we observed larger effect sizes in CP than in EP. This result may reflect the lower symptom burden in EP, which, on average, was in the “mildly ill” category (Leuct et al., 2005). For example, as predictive models require sufficient variation in the sample to learn individual differences (Tejavibulya et al., 2022), the better prediction results in CP may be due to the broader distribution of scores (Figure S1). Similarly, group differences are easier to detect when symptoms are more severe. With mild symptomatology, EP neurobiology may be more similar to controls than CP (Ganella et al., 2017). Unfortunately, our sample sizes were too small to stratify groups by symptom burden (e.g., CP with lower symptom burden) to test this consideration. Alternatively, a more speculative proposal is that symptom-based networks’ increased ‘aberrantness’ during the proposed transition from EP to CP could contribute to increased effect sizes. If this explanation is true, then changes in the anatomy of the symptom models could be targets for early interventions that minimize disease progression. Still, dense longitudinal and neuromodulation studies would be needed to test progression in these networks and how they associate with symptom change.

In EP, we observed that specific positive (but not negative) symptoms were more informative for prediction than others. This may be because the positive symptom brain biology stabilizes faster than negative symptoms. Another possible reason is the temporal worsening of the negative symptoms. Negative symptoms, if primary (Correll and Schooler, 2020), tend to become more severe throughout the psychosis lifetime, while positive symptoms can emerge once and remain dormant or happen again (George et al., 2017, Mosolov and Yaltonskaya, 2021). Unfortunately, we could not test this in CP because we lacked item-level information. Future studies with access to such data should investigate if similar patterns exist in CP.

Our study also has several limitations. First, we lacked a cohort with longitudinal data tracking the progression from first-episode psychosis to chronic psychosis. This data would be needed to support deeper inferences about causality, trajectories, or mechanisms. Further, cohorts with direct experimental manipulation (such as neuromodulation or randomized control trials) would strengthen such inferences. Second, we also lack medication information for CP, which may affect prediction performance and group differences. However, in EP, controlling for medication did not impact results. Third, EP and CP were from different unharmonized studies. Similarly, the datasets have many differences (i.e., dataset shifts). Thus, comparing these groups directly to understand disease progression is not possible. However, the symptom networks are similar across cohorts, and one model generalized. These results suggest a high level of generalizability despite significant dataset shifts. Finally, the model performance is still far below that of clinically actionable, which, if achievable, could take shape by providing therapeutic target sites, identifying EP treatment responsiveness characteristics, and—more ambitiously—predicting conversion from the prodrome and EP stages to the CP stage. Additional modalities or methods to characterize functional data are needed to improve model performance. Task-based connectivity (Jiang et al., 2020) and naturalistic paradigms such as movie watching (Finn et al., 2018) have been proven to predict better than resting-state data. Dynamic fMRI measures (Lurie et al., 2020; Ye et al., 2023) can also explain more psychiatric neurobiology than resting-state and static measures.

In the presented study, we examine the relationship between resting-state functional connectivity and psychosis symptom dimensions in EP and CP. Our results may provide a neurobiological reference point to temporally track edge-level, stage-related changes in functional connectivity related to symptom severity in psychosis populations. They also offer an investigational starting point for reconciling treatment response success in EP vs CP. To explore these questions, future studies should incorporate longitudinal data to more accurately evaluate symptom-based networks over time and neuromodulation to understand their causal effects on symptom changes. In addition, a treatment responsiveness metric should be included as a potential mediating variable for the identified connectivity changes.

## Data Availability

Raw imaging and behavioral data are available through the Human Connectome Project for Early Psychosis (https://nda.nih.gov/general-query.html?q=query=featured-datasets:Connectomes%20Related%20to%20Human%20Disease) and Strategic Research Program for Brain Sciences (https://www.synapse.org/#!Synapse:syn22317078).

https://www.humanconnectome.org/study/human-connectome-project-for-early-psychosis

## Code and model availability

The main CPM analysis scripts can be found at https://github.com/YaleMRRC/CPM. BioImage Suite was used for analysis and visualization (www.bisweb.yale.edu). Models are shared at https://www.nitrc.org/projects/bioimagesuite/.

## Data availability

Data are available through the Human Connectome Project Early Psychosis (https://www.humanconnectome.org/study/human-connectome-project-for-early-psychosis/data-releases) and the Strategic Research Program for Brain Multi-disorder Connectivity Dataset (https://bicr-resource.atr.jp/srpbsopen/).

## Author Contributions

MF and DS conceived the current study. JY processed some of the chronic psychosis data used in the study. MF preprocessed all early psychosis data and the majority of the chronic psychosis data. MF performed all data analysis. AP offered analysis and study design feedback. MF and DS wrote the paper. MF, AP, and DS discussed the results and their implications. MF, JY, AP, ND, and DS all edited and approved the paper.

## Acknowledgements

We thank the participants who contributed to the datasets we used in the study and the individuals involved in the data collection.

## Funding

M.F. was supported by the Quad Fellowship. J.Y. is supported by the Gruber Science Fellowship. We would like to also thank Brendan Adkinson for helpful feedback.

## Disclosures

None of the authors have any disclosures concerning the content of this manuscript.

## Notes

### Competing Interest Statement

The authors have declared no competing interest.

### Funding Statement

This study did not directly receive any funding. M.F. was supported by the Gruber and Quad Fellowships. J.Y. was supported by the Gruber Fellowship. Any findings, conclusions, or recommendations expressed in this material are those solely of the authors and do not necessarily reflect those of the funding agencies.

### Author Declarations

This study only used open-source human data originally located at the Human Connectome Project Early Psychosis and the Strategic Research Program for Brain Sciences.

### Summary of Updates

Minor revisions of the manuscript wording was done (strengthening the paper's argument) and additional citations were added.

