## Supplemental Materials for "Connectome-based predictive modeling of early and chronic psychosis symptoms" (from another version of this preprint: version 1, 2024-05-20)

**Supplementary Methods**

**Positive and Negative Syndrome Scale.** The Positive and Negative Syndrome Scale (PANSS) (Kay et al. 1987) measures dimensional psychosis symptoms. The PANSS assesses the presence and severity of positive, negative, and general psychopathology (GPS) within the past week through item-wise responses to questions specific to the positive, negative, or GPS symptom categories. The scale is widely used in clinical psychosis studies and has demonstrated reliability in assessing psychopathology across diverse patient populations (Lewandowski et al., 2020). The PANSS total, positive, negative, and general psychopathology scores were used as summary measures of psychosis severity. Summary measures are summations of itemized dimensions.

**Data preprocessing and data cleaning.** The acquisition and imaging parameters for the HCP-EP and SRPBS datasets are detailed elsewhere (Lewandowski et al. 2020, Tanaka et al., 2021). Preprocessing included motion correction, regressing mean white matter, cerebral spinal fluid, and gray matter time series, removing linear trends, and low-pass filtering. The Shen-268 parcellation (Shen et al. 2013) was used. First, the atlas was warped from MNI space into single-subject space. Next, the mean time course from each functionally defined node was calculated by averaging its constituent voxels’ time-series data. Pairwise correlations were computed between all node-pair combinations. These correlations were Fisher z-transformed to produce symmetric 268 × 268 connectivity matrices (Shen et al. 2017). Participants with an average imaging data frame-to-frame displacement exceeding 0.2 mm were excluded. The HCP-EP analysis was restricted to subjects with both anterior-posterior and posterior-anterior scans. Connectomes from the anterior-posterior and posterior-anterior scans with the lowest motion values were averaged to create an average connectome. For CP, missing nodes from any subject were removed from all participants (see Table S18) before CPM model training.

**EP Item-based Symptom Removal**. The HCP-EP dataset had individualized PANSS items (e.g., each symptom category could be further broken down into specific behavioral pathologies). To determine if a particular item was driving the prediction of a summary symptom score (positive, negative, total), we removed that item, re-ran CPM, and assessed the result. If prediction performance decreases, then that item drives prediction. If performance increases, then that item does not drive prediction.

**Connection length characterization**. We employ these standard approaches and summarize networks based on the length of connections (short- versus long-range connectivity). Euclidean distance between the centroids of each brain region in the Shen atlas was used to classify short and long-range edges. First, the distance between each pair of nodes was calculated as: √((𝑥0 −𝑥1)^2^ +(𝑦𝑜 −𝑦1)^2^ +(𝑧0 −𝑧1)^2^)), where (𝑥0,𝑦0,𝑧0), and (𝑥1𝑦1,𝑧1) represent the centroid for any two regions. Pairwise distances were separated into short, mid-short, mid-long, and long-distance connections based on quartiles. 2x4 contingency tables (Vassar Stats) were used to assess statistically significant distributions.

**Virtual lesion analysis.** CPM predictive networks are typically widespread and complex, so we conducted a virtual lesion analysis. For a CPM-based virtual lesion analysis, predictive networks can be set to zero to examine the degradation in predictive performance attributed to a virtual lesion of that network (Yip et al., 2020). We conducted two versions of virtual lesion analysis. First, we iteratively set each functional network to zero and examined how this impacted the model performance. We conducted this virtual lesion analysis for the canonical functional networks: medial frontal (MF), frontoparietal (FP), default mode (DMN), motor (MOT), visual I (VI), visual II (VII), visual association (VA), salience (SAL), subcortical (SC), and cerebellum (CBL). We used a leave-one-network-out approach, which entails removing a particular network from consideration during prediction training (network is set to zero) to examine how predictive performance changes due to the lesioning of that network. Iteratively, each functional network is set to zero and model performance was assessed. The impact of prediction was assessed with Steiger’s Z test (Steiger 1980, QuantPsy), which tests how two dependent correlations compare. In our case, we compared the median non-controlled model Pearsons’s r and the lesioned network model Pearsons’s r.

**Cross-Dataset Edge-Level Comparison**. Edge-level comparisons were measured with the *Matlab* hypergeometric cumulative (hygecdf) distribution function (<https://www.mathworks.com/help/stats/hygecdf.html>). The following input format was used to obtain a p-value: for two connectome comparisons c1 and c2, p = 1-*hygecdf*(# uniquely overlapping edges between c1 and c2, # uniquely significant edges in c1, # uniquely significant edges in c2, total # of unique edges possible in a node x node connectome). Let N=node. # unique edges = (N*N)-N/2. For example, with 268 nodes, the number of unique edges is 35778.

**Network-based statistics (NBS) for case-control group differences.** Case-control group differences in connectomes between EP or CP and HCs were conducted using a validated network-based statistical method with correction for multiple comparisons (Zalesky et al., 2010). Significance was assessed with a Z-score threshold of 1.98 and 1000 iterations.

**Supplementary Results**

**Impact of itemized symptom removal on EP CPM.** The positive model was lowered by removing the grandiosity and suspiciousness/persecution items, while removing hallucinations and delusions items enhanced prediction (Table S9). Specific items did not impact the total and negative score models (Table S9).

**Network Anatomy in EP.** Broadly, connections between the motor network and another network had the most edges in the model for both positive and negative edges. For the negative network, the cerebellum-motor internetwork had the highest number of contributions. Internetwork contributions with greater than 10 edges included the medial frontal network-cerebellum network, FP-CBL network, the motor-motor network, FP-motor network, motor-SC network, and the DMN-motor network. For the positive network, the distribution of contributive network-networks was broader. The motor-MF network had the highest contributions with the motor-VAs network coming in a close second. FPN connections with the MF and motor networks, as well as the motor and limbic network connection and limbic and subcortical network connections had contributions of 10 or more.

**Network Anatomy in CP.** Motor-other network connections dominated the predictive contributions in the positive and negative networks. For the negative network, internetwork connections with over 20 contributions included motor network connections with the motor network, limbic network, subcortical network. Connections over 10 included frontoparietal connections with the subcortical, motor, and medial frontal-cerebellum networks. Again, for the positive network, motor-other network connections were the most contributive, with 7/10 possible network connections over 10. The top five networks with the most contributions were the medial frontal, frontoparietal, motor, limbic, and subcortical networks. Other high connections include the medial frontal-default mode, visual association, and limbic networks.

**Supplementary Figures**

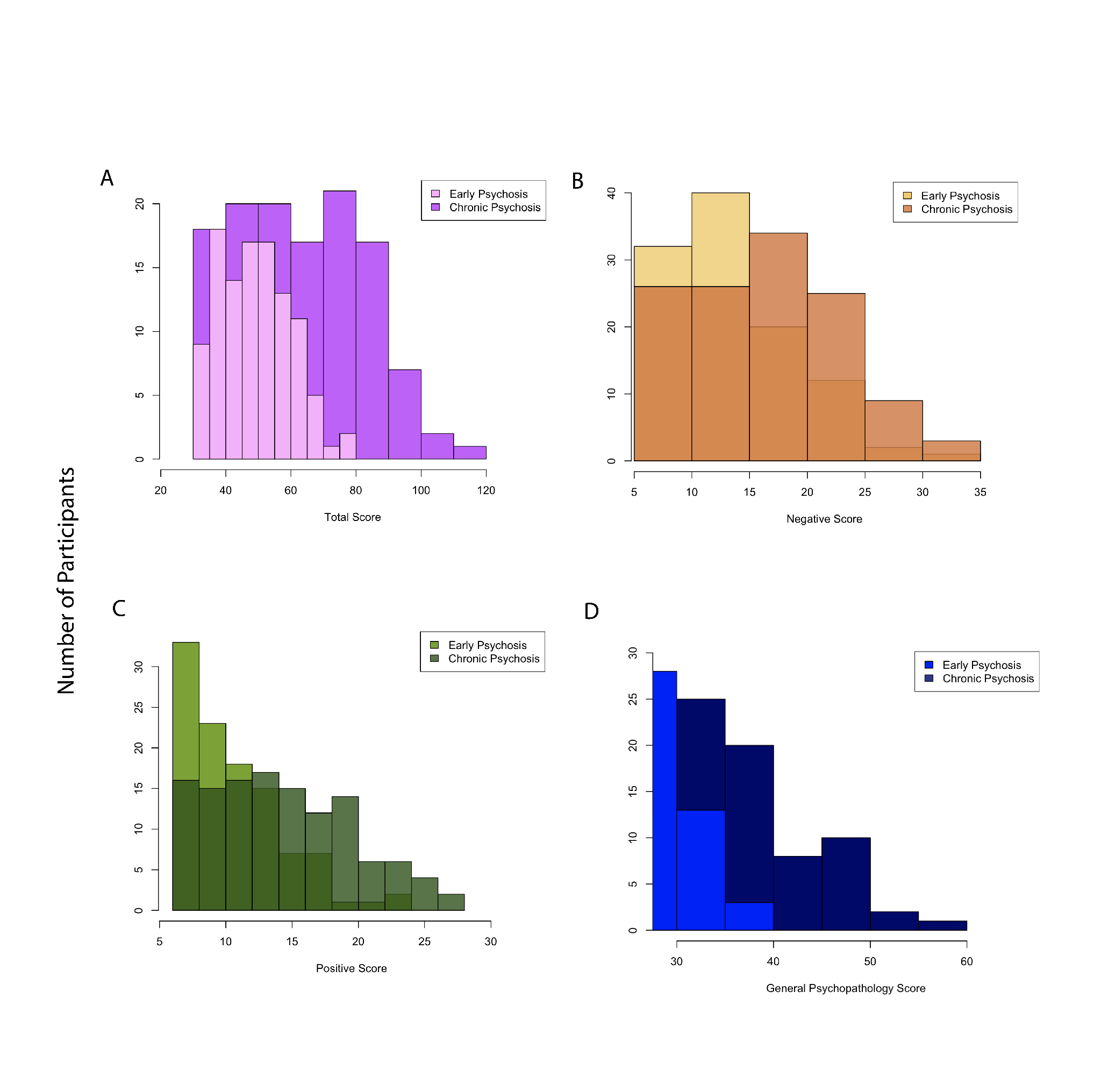

**Figure S1.** *PANSS Scores in CP and EP.* Histogram distributions of EP and CP PANSS symptom scores. A) Total symptoms. B) Negative symptoms. C) Positive symptoms. D) General psychopathology symptoms. EP = early psychosis, CP = chronic psychosis.

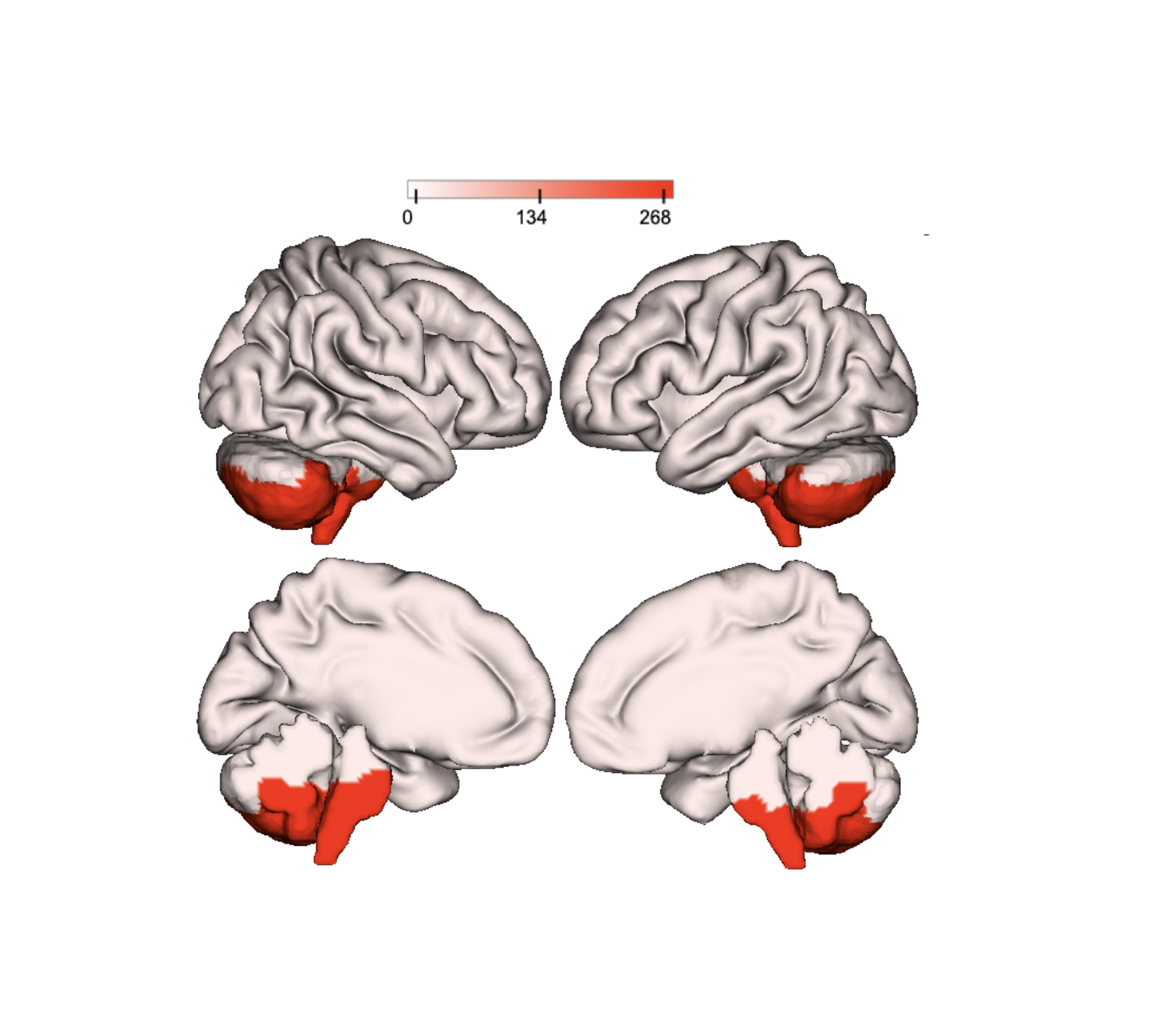

**Figure S2**. *Node-level elimination of zero-valued nodes.* During quality control for the SRPBS dataset, we excluded 26 brain nodes due to incomplete slice coverage. The spatial location of excluded nodes (in red) is predominantly in the cerebellum network. Missing nodes also belonged to the frontoparietal network (4 nodes), default mode network (2 nodes), motor network (1 node), 1st visual network (1 node), and the 2nd visual network (1 node).

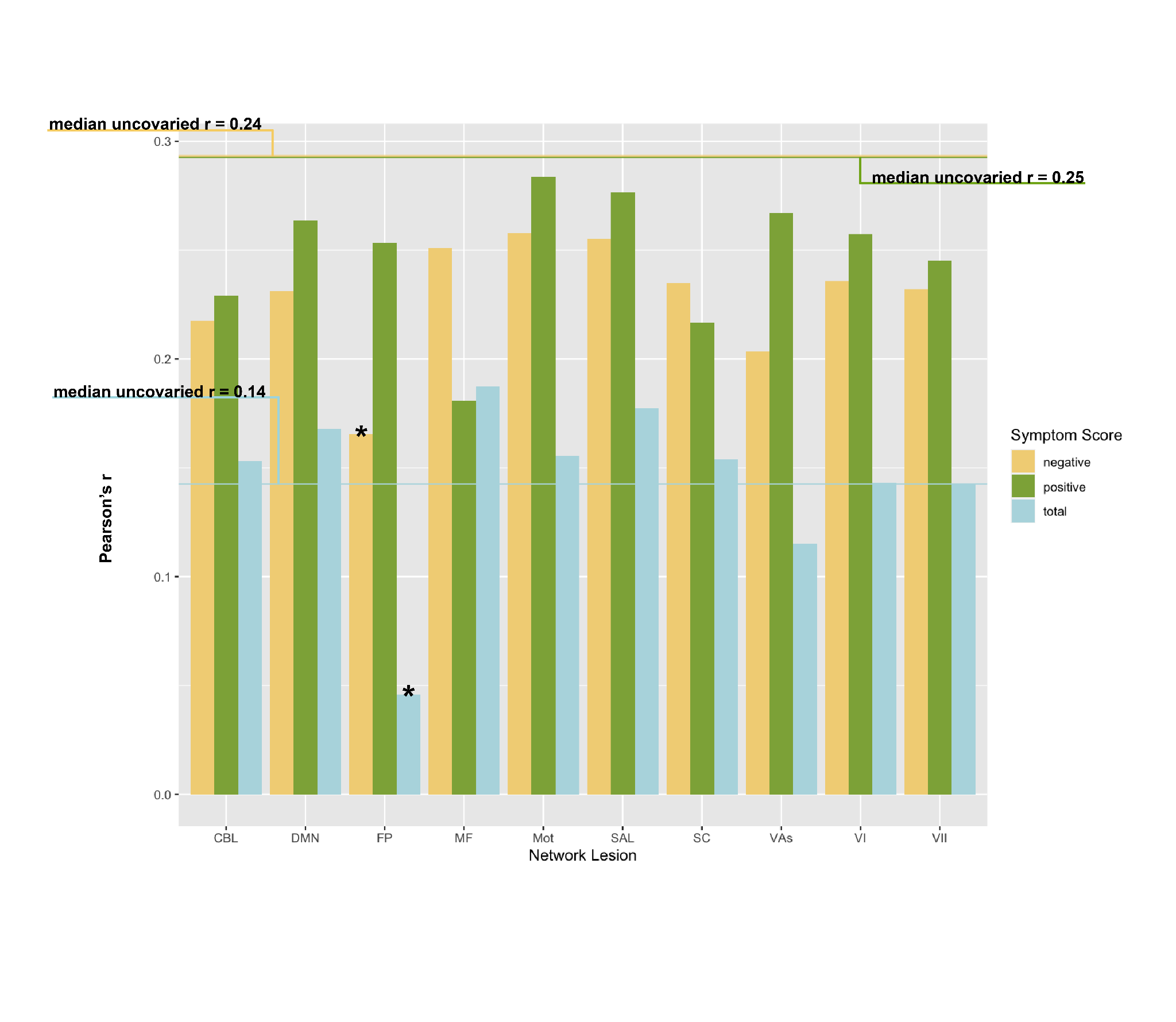

**Figure S3.** *Virtual Lesion Studies – Early Psychosis***.** Leave-one-out CPM results for each PANSS score in EP. Frontoparietal network lesioning during negative and total symptoms prediction significantly (*) impacted performance. Plotted values were calculated by iteratively eliminating each canonical network listed on the x-axis for each symptom condition. CBL=cerebellar network; DMN=default mode network; FP=frontal parietal network; MF=medial frontal network; Mot=motor-sensory network; SAL=salience network; SC=subcortical network; VAs=visual association network; V1=visual network 1; VII=visual network 2.

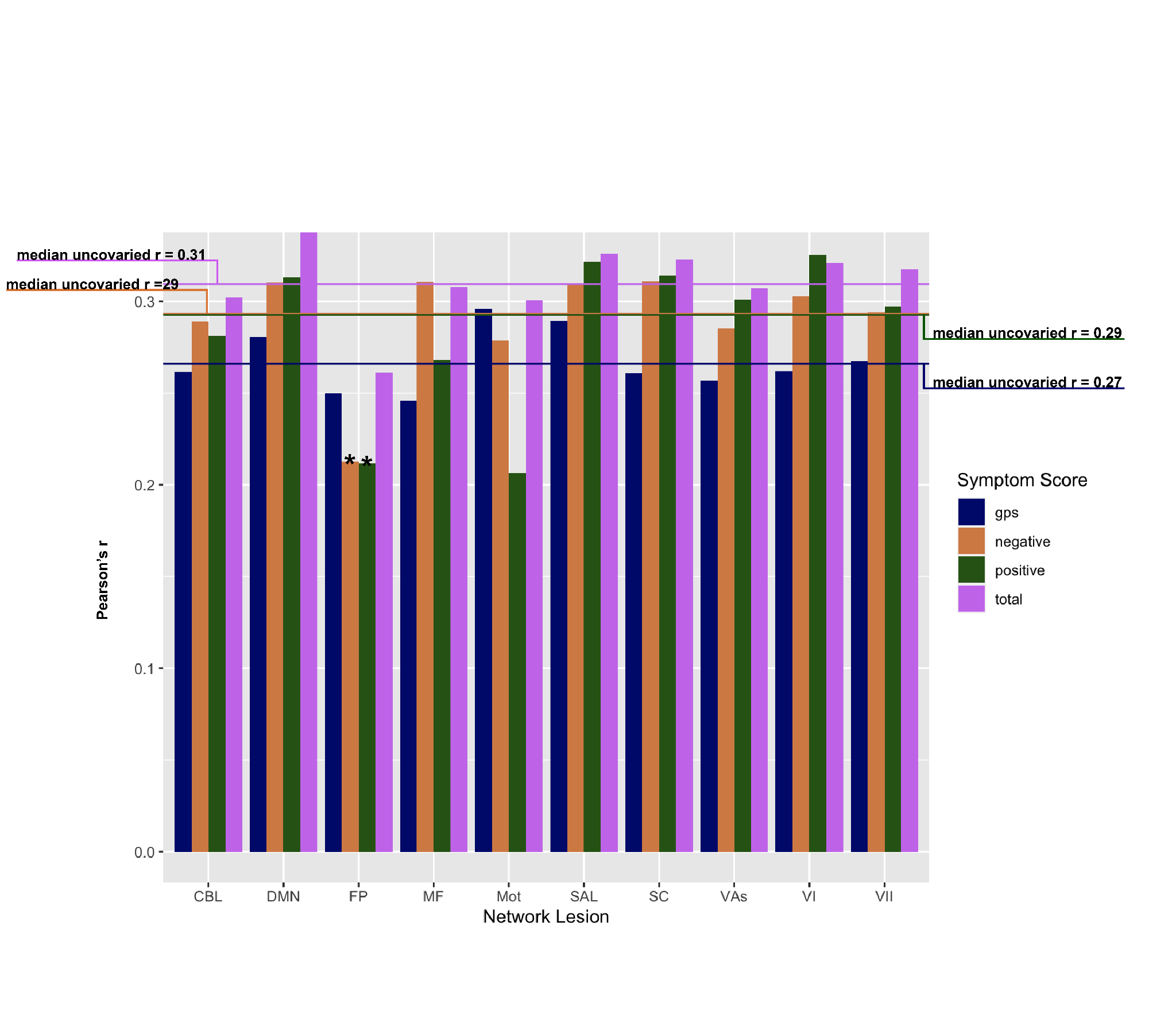

**Figure S4.** *Virtual Lesion Studies – Chronic Psychosis***.** Leave-one-out CPM results for each PANSS score in CP. Frontoparietal network lesioning during positive and negative symptoms prediction significantly (*=<0.05) impacted performance. Plotted values were calculated by iteratively eliminating each canonical network listed on the x-axis for each symptom condition. CBL=cerebellar network; DMN=default mode network; FP=frontal parietal network; MF=medial frontal network; Mot=motor-sensory network; SAL=salience network; SC=subcortical network; VAs=visual association network; V1=visual network #1; VII=visual network #2.

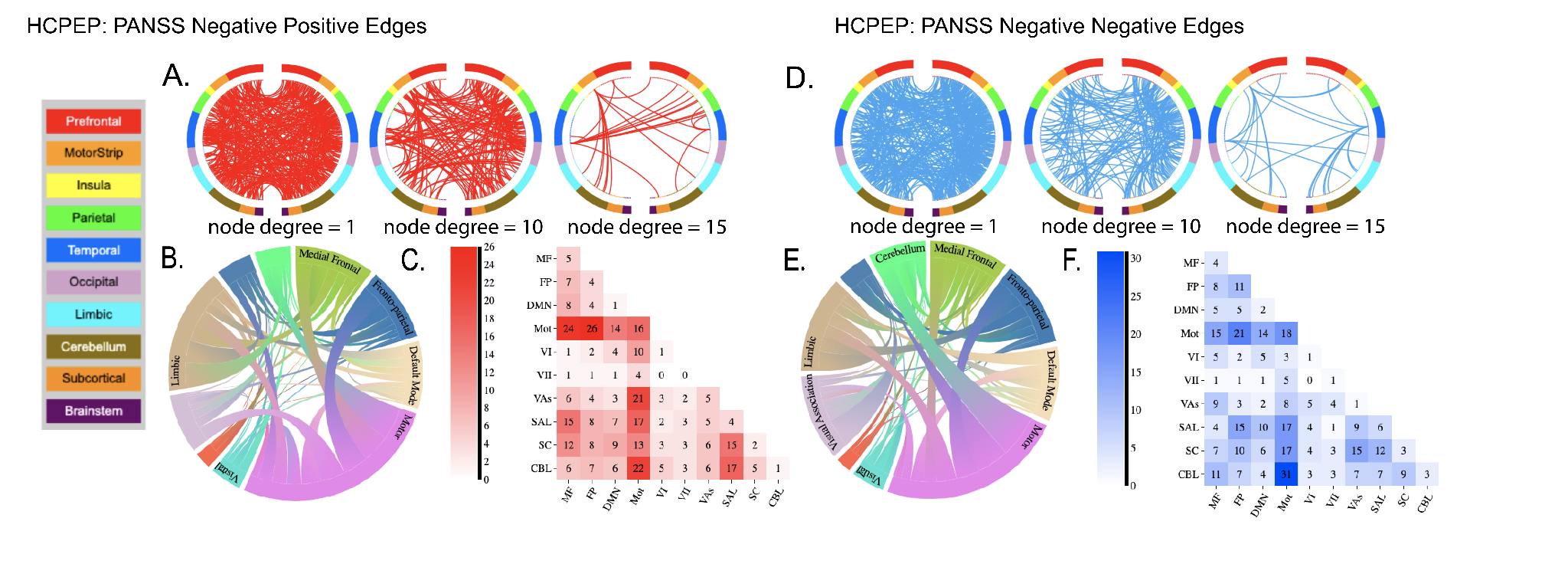

**Figure S5.** *Edge and Network-level contributions to predicting early negative symptoms scores*. Positive. (A) Interhemispheric and intrahemispheric network connections from positive networks that contribute to prediction of EP negative symptom scores. (B) Chord plot of network-level connectivity of the positive network that contributed to prediction of negative EP symptoms scores. (C) Network-level contribution from the positive network to EP negative symptoms score prediction. Negative. (D) Interhemispheric and intrahemispheric network connections from negative networks that contribute to prediction of EP negative symptom scores. (E) Chord plot of network-level connectivity of the negative network that contributed to prediction of negative EP symptoms scores. (F) Network-level contribution from the negative network to EP negative symptoms score prediction. EP = early psychosis, CP = chronic psychosis. The model visualized is a consensus model of the 10-cross-validated trials of the median performing train-test iteration.

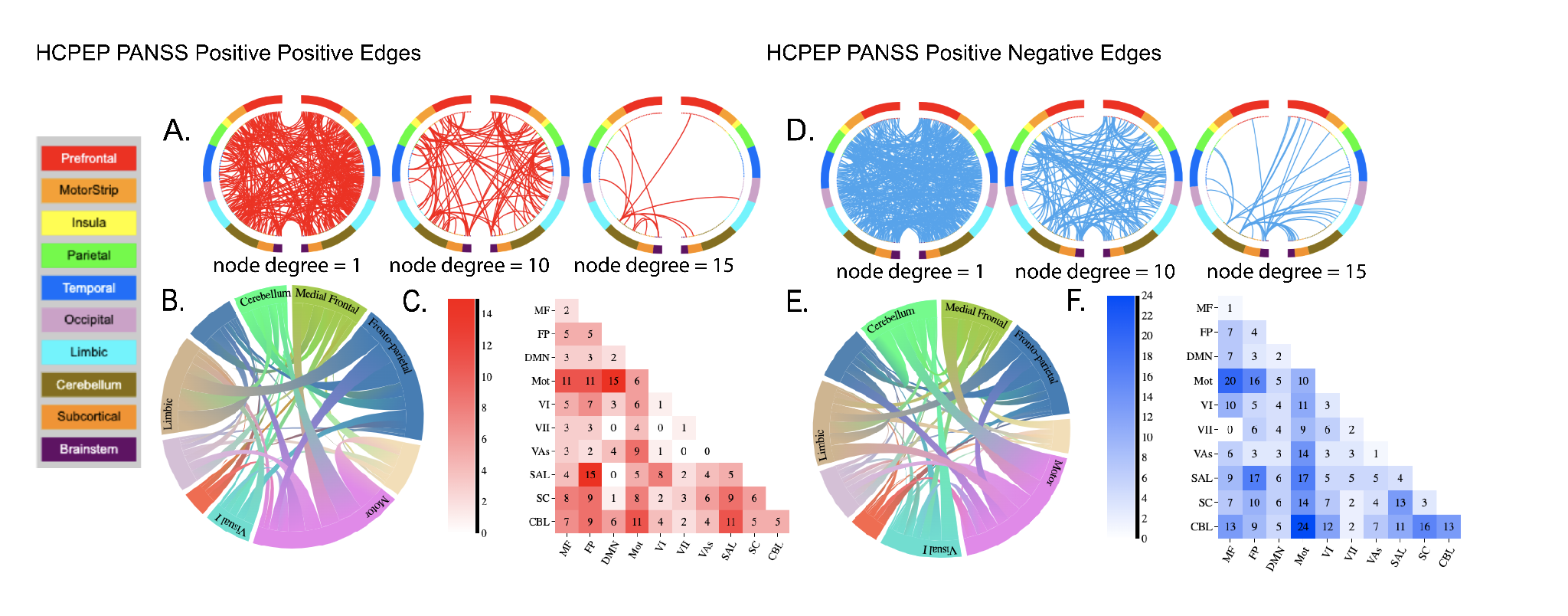

**­­**

**Figure S6.** *Edge and Network-level contributions to predicting early positive symptoms scores*. Positive. (A) Interhemispheric and intrahemispheric network connections from positive networks that contribute to prediction of EP positive symptom scores. (B) Chord plot of network-level connectivity of the positive network that contributed to the prediction of EP positive symptom scores. (C) Network-level contribution from the positive network to EP positive symptoms score prediction. Negative. (D) Interhemispheric and intrahemispheric network connections from the negative network that contribute to prediction of EP positive symptom scores. (E) Chord plot of network-level connectivity of the negative network that contributed to prediction of EP positive symptoms scores. (F) Network-level contribution from the negative network to EP positive symptoms score prediction. EP = early psychosis, CP = chronic psychosis. The model visualized is a consensus model of the 10-cross-validated trials of the median performing train-test iteration.

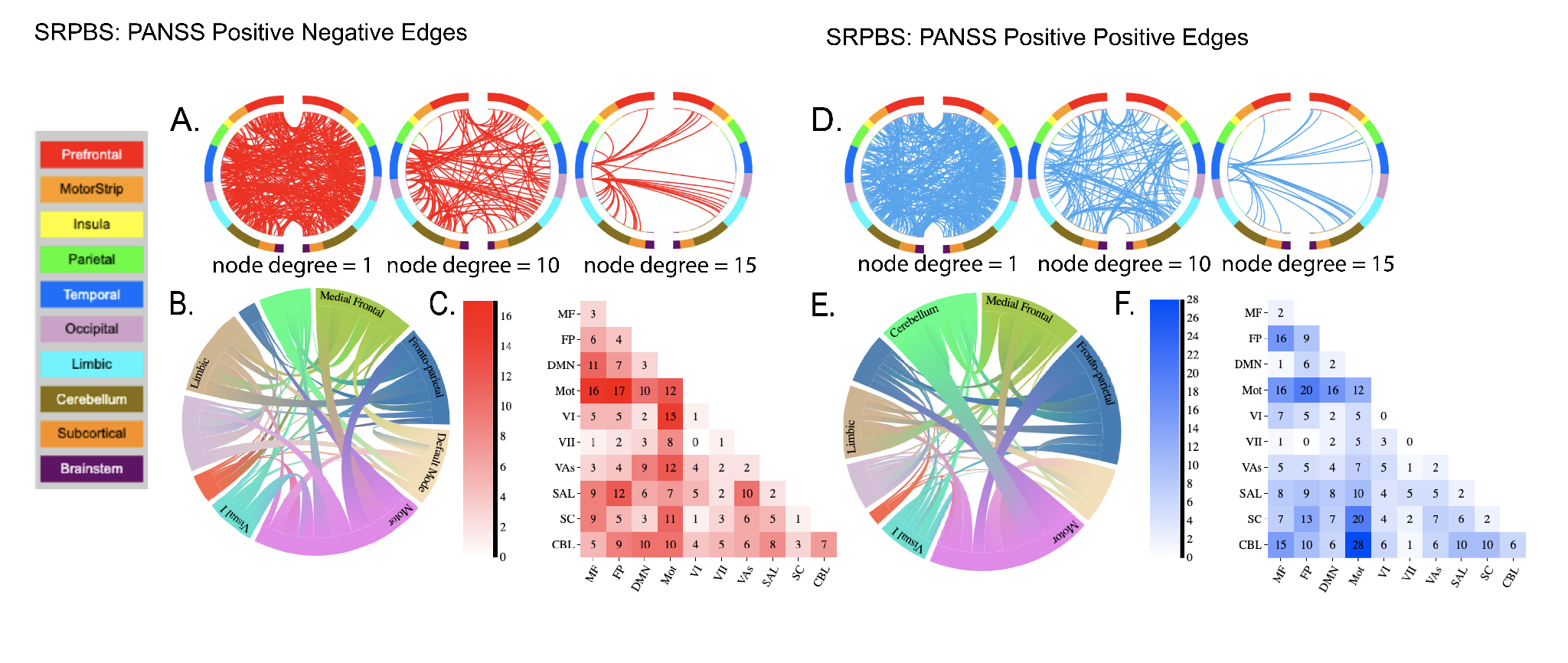

**Figure S7.** *Edge and Network-level contributions to predicting chronic positive symptoms scores.* Positive. (A) Interhemispheric and intrahemispheric network connections from positive networks that contribute to prediction of CP positive symptom scores. (B) Chord plot of network-level connectivity of the positive network that contributed to prediction of CP positive symptoms scores. (C) Network-level contribution from the positive network to CP positive symptoms score prediction. Negative. (D) Interhemispheric and intrahemispheric network connections from negative networks that contribute to prediction of CP positive symptom scores. (E) Chord plot of network-level connectivity of the negative network that contributed to prediction of CP positive symptoms scores. (F) Network-level contribution from the negative network to CP positive symptoms score prediction. EP = early psychosis, CP = chronic psychosis. The model visualized is a consensus model of the 10-cross-validated trials of the median performing train-test iteration.

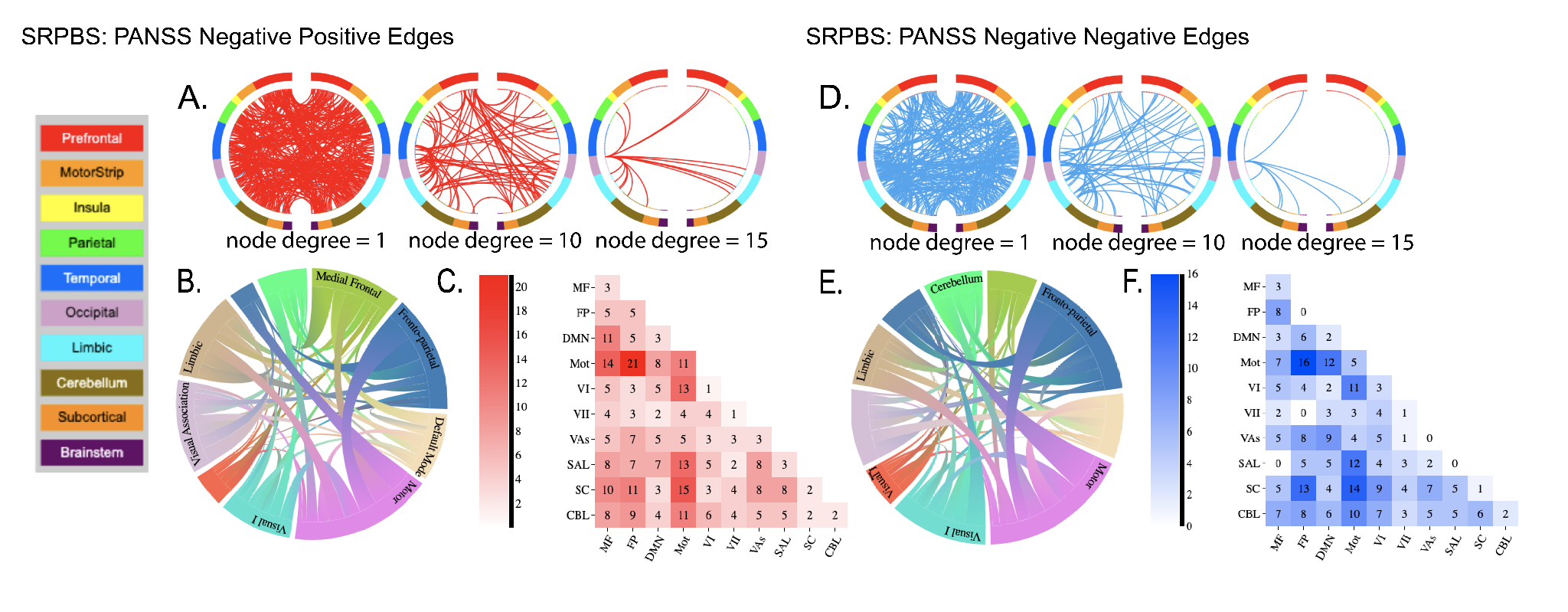

**Figure S9.** *Edge and Network-level contributions to predicting chronic negative symptoms scores*. Positive. (A) Interhemispheric and intrahemispheric network connections from the positive network that contributed to prediction of CP negative symptom scores. (B) Chord plot of network-level connectivity of the positive network that contributed to prediction of CP negative symptoms scores. (C) Network-level contribution from the positive network to CP negative symptoms score prediction. Negative. (D) Interhemispheric and intrahemispheric network connections from the negative network that contributed to prediction of CP negative symptom scores. (E). Chord plot of network-level connectivity of the negative network that contributed to prediction of CP negative symptoms scores. (F). Network-level contribution from the negative network to CP negative symptoms score prediction. EP = early psychosis, CP = chronic psychosis. The model visualized is a consensus model of the 10-cross-validated trials of the median performing train-test iteration.

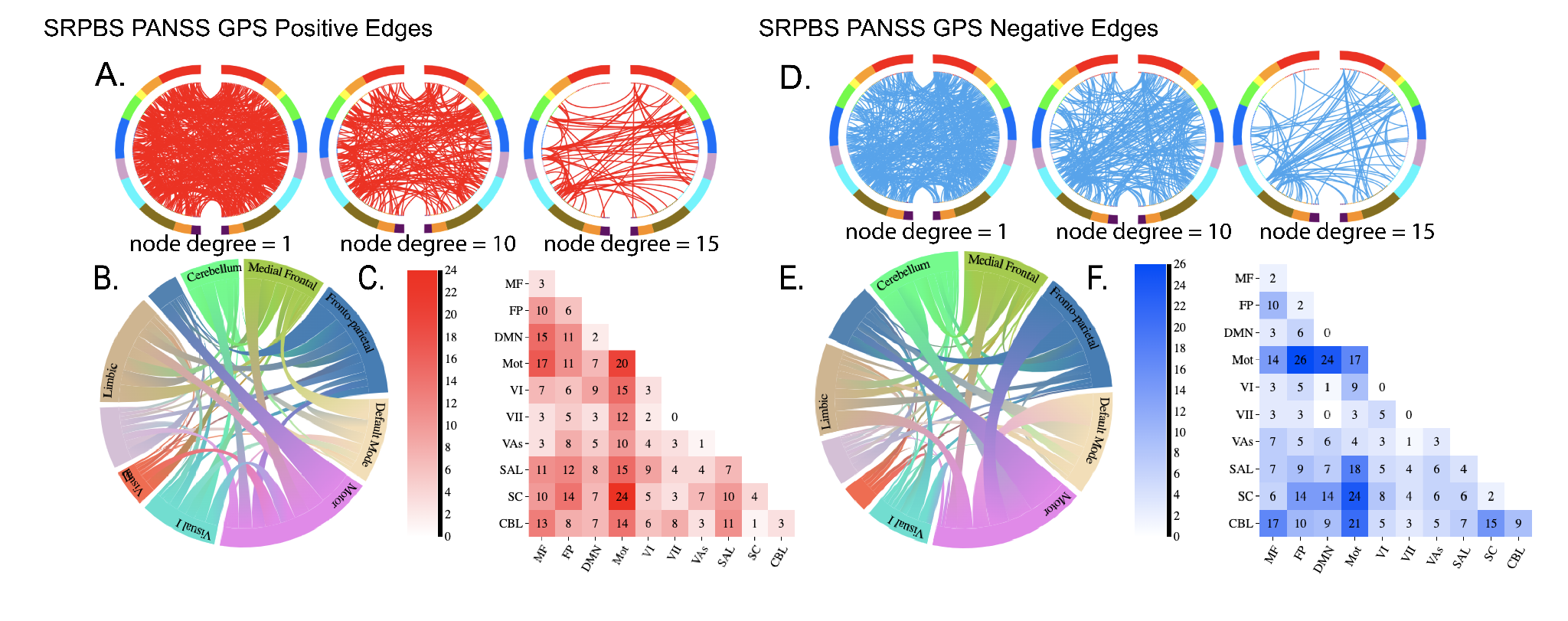

**Figure S10**. *Edge and Network-level contributions to predicting chronic general psychopathology symptoms scores*. Positive. (A) Interhemispheric and intrahemispheric network connections from the positive network that contributed to prediction of CP general psychopathology symptom scores. (B) Chord plot of network-level connectivity of the positive network that contributed to prediction of CP general psychopathology symptoms scores. (C) Network-level contribution from the positive network to CP general psychopathology symptoms score prediction. Negative. (D) Interhemispheric and intrahemispheric network connections from the negative network that contributed to prediction of CP general psychopathology symptom scores. (E) Chord plot of network-level connectivity of the negative network that contributed to prediction of CP general psychopathology symptoms scores. (F) Network-level contribution from the negative network to CP general psychopathology symptoms score prediction. EP = early psychosis, CP = chronic psychosis. The model visualized is a consensus model of the 10-cross-validated trials of the median performing train-test iteration.

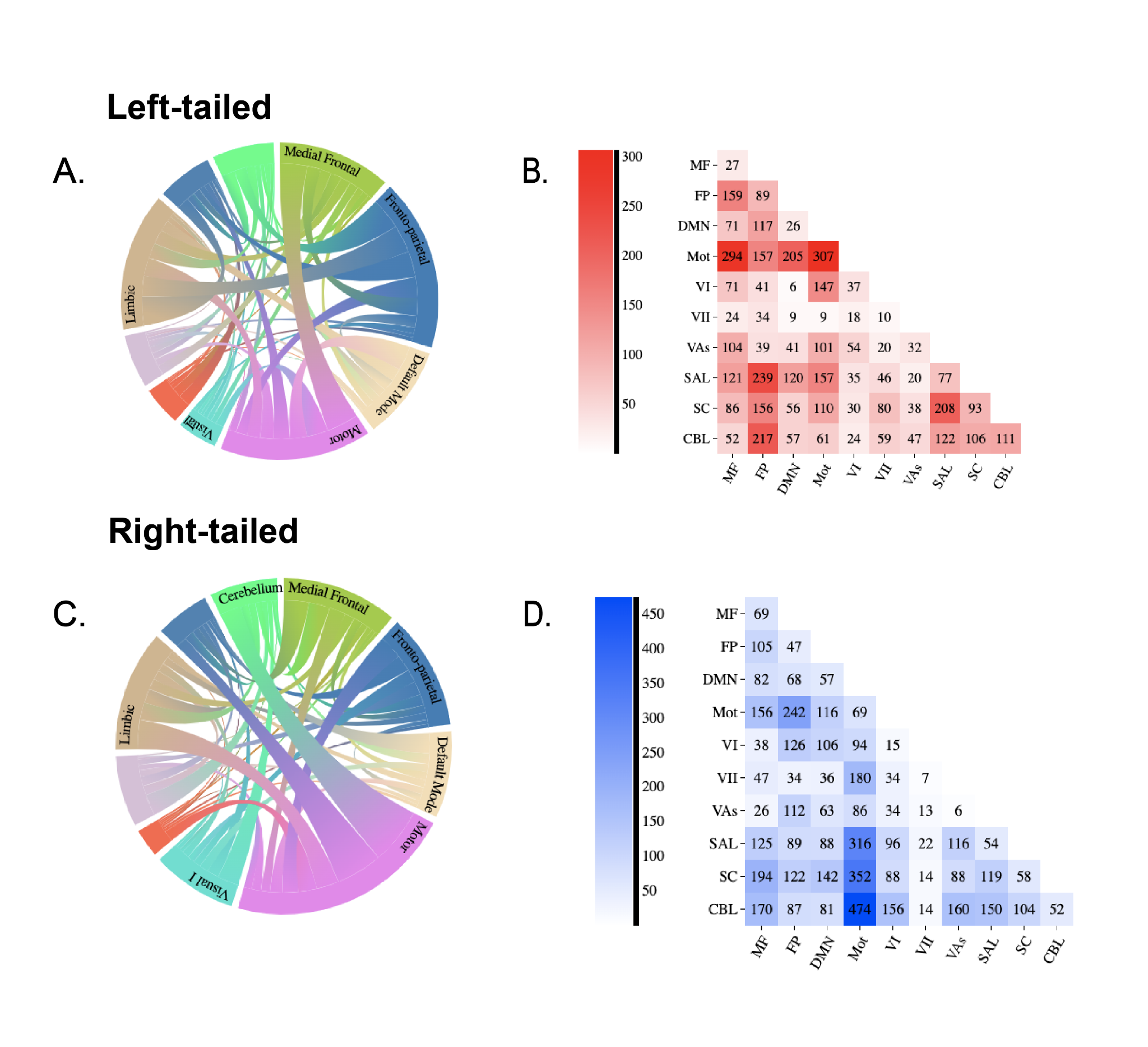

**Figure S11.** *Group Differences: Chronic Psychosis versus Healthy Controls Network*. Left-tailed (HC < CP): (A) Chord plot of network-level connectivity differences in the left-tailed network between HC and CP. (B) Network-level differences between HC and CP in the left-tailed network. Right-tailed (HC > CP): (C) Chord plot of network-level connectivity differences in the right-tailed network between HC and CP. (D) Network-level differences between HC and CP in the right-tailed network. HC = healthy controls. EP = early psychosis, CP = chronic psychosis.

**Supplementary Tables**

| **Race** | **HCP-EP Patients** | **SRPBS** |
| --- | --- | --- |
| American Indian/Alaska Native | 1 | N/A |
| Asian | 7 | 123 |
| Black or African American | 40 | N/A |
| Multiracial | 1 | N/A |
| Unknown/Not Reported | 3 | N/A |
| White | 55 | N/A |

### **Table S1:** *Subject Race Distribution*. Racial make-up of early psychosis (EP) and chronic psychosis (CP) population. HCP-EP = Human Connectome Project Early Psychosis, SRPBS = Strategic Research Program for Brain Sciences.

| **Site** | **Dataset** | **Scanner Type** | **Protocol** | **Sample Size** |
| --- | --- | --- | --- | --- |
| **Brigham and Women's Hospital** | **HCP-EP** | Siemens MAGNETOM Prisma 3T | Multiband sequence and 32 channel head coil | 20 |
| **Indiana University** | **HCP-EP** | Siemens MAGNETOM Prisma 3T | Multiband sequence and 32 channel head coil | 54 |
| **Massachusetts General Hospital** | **HCP-EP** | Siemens MAGNETOM Prisma 3T | Multiband sequence and 64 channel head coil | 8 |
| **McLean University** | **HCP-EP** | Siemens MAGNETOM Prisma 3T | Multiband sequence and 64 channel head coil | 25 |
| **Kyoto University** | **SRPBS** | Siemens TimTrio 3T | Siemens ep2d_bold sequence and 32 channel head coil | 82 |
| **Showa University** | **SRPBS** | Siemens Verio 3T | Siemens ep2d_bold sequence and 12 channel head coil | 8 |
| **University of Tokyo Hospital** | **SRPBS** | General Electric MR750W 3T | Siemens ep2d_bold sequence equivalent and 24 channel head coil | 33 |

**Table S2:** *Site-Specific Scanner Information***.** Imaging proctors and scanner characteristics are reported for each site represented in the study. HCP-EP = Human Connectome Project Early Psychosis, SRPBS = Strategic Research Program for Brain Sciences.

|  | **Covariate** | **r*** | **MSE*** | **p-value** |
| --- | --- | --- | --- | --- |
| PANSS Positive |  | | | |
|  | None | 0.2506 | 14.1760 | 0.01 |
|  | Age | 0.2550 | 14.3255 | 0.01 |
|  | Medication Exp | 0.2378 | 14.5877 | 0.01 |
|  | Motion | 0.2242 | 14.6632 | 0.01 |
|  | Phenotype | 0.2375 | 14.4126 | 0.01 |
|  | Sex | 0.2523 | 13.4093 | 0.01 |
|  | Site | 0.2035 | 14.8495 | 0.01 |
| PANSS Negative |  | | | |
|  | None | 0.2418 | 29.6276 | 0.01 |
|  | Age | 0.2234 | 30.1741 | 0.01 |
|  | Medication Exp | 0.2542 | 28.9869 | 0.01 |
|  | Motion | 0.2058 | 30.4668 | 0.01 |
|  | Phenotype | 0.2153 | 30.3156 | 0.01 |
|  | Sex | 0.2209 | 30.1067 | 0.01 |
|  | Site | 0.1955 | 30.5429 | 0.01 |
| PANSS  GPS | None | -0.0111 | 33.9235 | 0.12 |
|  | Age | -0.0046 | 35.1862 | 0.02 |
|  | Medication Exp | 0.0312 | 32.8754 | 0.01 |
|  | Motion | 0.0224 | 32.9628 | 0.06 |
|  | Phenotype | -0.0076 | 33.4499 | 0.08 |
|  | Sex | -0.0133 | 34.3126 | 0.03 |
|  | Site | 0.0423 | 33.2017 | 0.03 |
| PANSS  Total | None | 0.1426 | 127.1850 | 0.01 |
|  | Age | 0.1597 | 125.0778 | 0.01 |
|  | Medication Exp | 0.1743 | 123.9503 | 0.01 |
|  | Motion | 0.1418 | 127.9444 | 0.01 |
|  | Phenotype | 0.1074 | 131.3769 | 0.01 |
|  | Sex | 0.1326 | 129.5790 | 0.01 |
|  | Site | 0.1419 | 126.8260 | 0.01 |

**Table S3:** *PANSS Score Prediction performance for the median model in early psychosis disorder***.** CPM results while controlling for key EP covariates for all PANSS summary measures. *Pearson’s correlation (r) and mean square error (MSE) between observed and predicted values

| **PANSS Score** | **Network Lesion** | **Symptom Score Variance** | **r*** | **MSE*** | **p-value** | **Z score** | **1-tailed p** |
| --- | --- | --- | --- | --- | --- | --- | --- |
| Positive | Delusions | 1.628 | 0.2751 | 8.2647 | 0.01 | -1.055 | 0.1458 |
|  | Conceptual Disorganization | 0.807 | 0.2327 | 13.1313 | 0.01 | 0.492 | 0.3115 |
|  | Hallucinations | 2.229 | 0.2896 | 10.2234 | 0.01 | -0.758 | 0.2242 |
|  | Excitement | 0.402 | 0.2442 | 13.2894 | 0.01 | 0.203 | 0.4198 |
|  | Grandiosity | 1.386 | 0.1997 | 11.0675 | 0.01 | 1.443 | 0.0746 |
|  | Suspiciousness/persecution | 1.581 | 0.1854 | 10.3145 | 0.01 | 1.664 | 0.0481 |
|  | Hostility | 0.479 | 0.2442 | 12.717 | 0.01 | 0.161 | 0.4362 |
|  | Hallucinations and Delusions | 5.239 | 0.337 | 5.3645 | 0.01 | -2.205 | 0.01374 |
| Negative |  | | | | | |  |
|  | Blunted Affect | 2.015 | 0.221 | 19.1438 | 0.01 | 0.482 | 0.3148 |
|  | Emotional Withdrawal | 1.458 | 0.238 | 21.5015 | 0.01 | 0.086 | 0.4656 |
|  | Poor Rapport | 0.67 | 0.219 | 25.6515 | 0.01 | 0.538 | 0.2953 |
|  | Passive/Apathetic Social Withdrawal | 1.444 | 0.229 | 21.6431 | 0.01 | 0.351 | 0.3629 |
|  | Difficulty in Abstract Thinking | 1.612 | 0.240 | 23.159 | 0.01 | 0.048 | 0.4811 |
|  | Lack of Spontaneity and Flow of Conversation | 1.641 | 0.237 | 20.0229 | 0.02 | 0.14 | 0.4442 |
|  | Stereotyped Thinking | 0.547 | 0.224 | 26.4738 | 0.01 | 0.457 | 0.3237 |
| Total | Positive Symptoms | 14.527 | 0.125 | 88.1956 | 0.01 | 0.281 | 0.3892 |
|  | Negative Symptoms | 30.170 | 0.140 | 66.403 | 0.01 | 0.049 | 0.4803 |
|  | General Psychopathology Symptoms | 51.583 | 0.187 | 51.1187 | 0.01 | -0.931 | 0.1760 |

### **Table S4:** *Itemized symptom removal prediction performance for median model in early psychosis disorder***.** Itemized symptom removal helped determine which symptom-category items CPM was partial to predicting.

| **Dataset** | **PANSS Score** | **Edge sign** | **L** | **R** | **Interhemispheric** | **Total** | **Chi squared (*,**)** |
| --- | --- | --- | --- | --- | --- | --- | --- |
| SRPBS | Total | Positive | 102 | 206 | 450 | 758 | 10.57 (1,0.001) |
|  |  | Negative | 166 | 156 | 362 | 684 | 0.05 (1,0.823) |
|  | Positive | Positive | 128 | 188 | 348 | 664 | 3.35 (1,0.067) |
|  |  | Negative | 212 | 168 | 372 | 752 | 1.45 (1,0.229) |
|  | Negative | Positive | 96 | 186 | 388 | 670 | 8.62 (1,0.003) |
|  |  | Negative | 138 | 132 | 298 | 568 | 0.01 (1,0.920) |
|  | General Psychopathology | Positive | 172 | 204 | 482 | 858 | 0.71 (1,0.399) |
|  |  | Negative | 224 | 194 | 402 | 820 | 0.57 (1,0.450) |
| HCP-EP |  | | | | | | |
|  | Total | Positive | 152 | 170 | 302 | 624 | 0.23 (1,0.632) |
|  |  | Negative | 176 | 156 | 366 | 698 | 0.28 (1,0.597) |
|  | Positive | Positive | 204 | 144 | 200 | 548 | 3.14 (1,0.076) |
|  |  | Negative | 202 | 164 | 462 | 828 | 1.07 (1,0.301) |
|  | Negative | Positive | 240 | 166 | 370 | 776 | 4.11 (1,0.043) |
|  |  | Negative | 284 | 154 | 338 | 776 | 12.43 (1,0.0004) |

**Table S5:** *Hemispheric Connections*. Statistical evaluation of hemispheric connection profiles in EP and CP symptom networks. Connection-type count and chi-squared values are reported. *p values, **degrees of freedom, L=left, R=right. HCP-EP = Human Connectome Project Early Psychosis, SRPBS = Strategic Research Program for Brain Sciences.

| **Dataset** | **PANSS Score** | **Edge sign** | **Short** | **Mid-Short** | **Mid-Long** | **Long** | **Total** | **Chi squared (*,**)** |
| --- | --- | --- | --- | --- | --- | --- | --- | --- |
| SRPBS | Total | Positive | 174 | 208 | 210 | 166 | 758 | 0.59  (1,0.442) |
|  |  | Negative | 158 | 206 | 168 | 152 | 684 | 1.33  (1,0.249) |
|  | Positive | Positive | 180 | 134 | 194 | 156 | 664 | 1.41  (1,0.235) |
|  |  | Negative | 168 | 184 | 214 | 186 | 752 | 0.1 (1,0.752) |
|  | Negative | Positive | 160 | 170 | 172 | 168 | 670 | 0.03  (1,0.863) |
|  |  | Negative | 162 | 140 | 152 | 114 | 568 | 0.27  (1,0.603) |
|  | GPS | Positive | 198 | 238 | 232 | 190 | 858 | 0.75  (1,0.387) |
|  |  | Negative | 216 | 234 | 178 | 192 | 820 | 0.11  (1,0.740) |
| HCP-EP |  | | | | | | | |
|  | Total | Positive | 154 | 142 | 190 | 138 | 624 | 0.05  (1,0.823) |
|  |  | Negative | 166 | 134 | 196 | 202 | 698 | 0.65  (1,0.420) |
|  | Positive | Positive | 166 | 138 | 124 | 120 | 548 | 0.47  (1,0.493) |
|  |  | Negative | 184 | 210 | 250 | 184 | 828 | 0.31  (1,0.578) |
|  | Negative | Positive | 166 | 220 | 216 | 174 | 776 | 1.63  (1,0.202) |
|  |  | Negative | 202 | 174 | 204 | 196 | 776 | 0.38  (1,0.538) |

**Table S6:** *Connection Ranges*. Statistical evaluation of connection range profiles in EP and CP symptom networks. Connection-type count and chi-squared values are reported. *p values in parentheses, GPS = general psychopathology. HCP-EP = Human Connectome Project Early Psychosis, SRPBS = Strategic Research Program for Brain Sciences.

| **PANSS Score** | **Network** | **r*** | **MSE*** | **p-value** | **Z score** | **1-tailed p** |
| --- | --- | --- | --- | --- | --- | --- |
| **Positive** | Medial Frontal | 0.1809 | 14.87 | < 0.0001 | 1.289 | 0.10 |
|  | Frontoparietal | 0.2534 | 14.25 | < 0.0001 | -0.077 | 0.47 |
|  | Default Mode | 0.2635 | 14.02 | < 0.0001 | -0.4 | 0.35 |
|  | Motor | 0.2838 | 13.94 | < 0.0001 | -1.007 | 0.16 |
|  | Visual I | 0.2573 | 14.31 | < 0.0001 | -0.21 | 0.42 |
|  | Visual II | 0.245 | 14.48 | < 0.0001 | 0.171 | 0.43 |
|  | Visual Association | 0.2669 | 14.06 | < 0.0001 | -0.458 | 0.32 |
|  | Limbic | 0.2766 | 13.91 | < 0.0001 | -0.886 | 0.19 |
|  | Subcortical | 0.2168 | 15.01 | < 0.0001 | 0.839 | 0.20 |
|  | Cerebellum | 0.2291 | 14.82 | < 0.0001 | 0.662 | 0.25 |
| **Negative** | Medial Frontal | 0.2511 | 29.23 | 0.01 | -0.255 | 0.40 |
|  | Frontoparietal | 0.1660 | 31.845 | 0.02 | 1.815 | 0.04 |
|  | Default Mode | 0.2358 | 29.68 | 0.01 | 0.4318 | 0.43 |
|  | Motor | 0.2546 | 29.60 | 0.01 | -0.26 | 0.40 |
|  | Visual I | 0.2359 | 29.74 | 0.01 | 0.133 | 0.45 |
|  | Visual II | 0.2321 | 29.21 | 0.04 | 0.22 | 0.41 |
|  | Visual Association | 0.2034 | 30.46 | 0.02 | 0.86 | 0.19 |
|  | Limbic | 0.2552 | 28.95 | 0.02 | -0.32 | 0.37 |
|  | Subcortical | 0.2399 | 29.63 | 0.01 | 0.043 | 0.48 |
|  | Cerebellum | 0.226 | 29.77 | 0.01 | 0.363212 | 0.73 |
| **Total** | Medial Frontal | 0.1873 | 121.84 | < 0.0001 | -0.918 | 0.18 |
|  | Frontoparietal | 0.0458 | 137.61 | < 0.0001 | 1.674 | 0.05 |
|  | Default Mode | 0.1678 | 125.807 | < 0.0001 | -0.625 | 0.27 |
|  | Motor | 0.1556 | 129.32 | < 0.0001 | -0.104 | 0.46 |
|  | Visual I | 0.1431 | 128.72 | < 0.0001 | -0.01 | 0.50 |
|  | Visual II | 0.1429 | 127.61 | < 0.0001 | -0.007 | 0.50 |
|  | Visual Association | 0.1152 | 132.33 | < 0.0001 | 0.532 | 0.30 |
|  | Limbic | 0.1775 | 124.09 | < 0.0001 | -0.703 | 0.24 |
|  | Subcortical | 0.1539 | 128.12 | < 0.0001 | -0.227 | 0.41 |
|  | Cerebellum | 0.1529 | 129.16 | < 0.0001 | -0.24 | 0.41 |

### **Table S7:** *Virtual lesion prediction performance for median model in early psychosis disorder*.

| **PANSS Score** | **Covariate** | **r*** | **MSE*** | **p-value** |
| --- | --- | --- | --- | --- |
| **Positive** |  | | | |
|  | None | 0.2926 | 26.1426 | 0.01 |
|  | Age | 0.2904 | 25.9032 | 0.04 |
|  | Motion | 0.2985 | 26.0264 | 0.01 |
|  | Sex | 0.2661 | 26.6064 | 0.02 |
|  | Site | 0.3068 | 25.7044 | 0.01 |
| **Negative** |  | | | |
|  | None | 0.2933 | 37.9273 | 0.02 |
|  | Age | 0.2981 | 38.1072 | 0.01 |
|  | Motion | 0.2943 | 37.8288 | 0.03 |
|  | Sex | 0.2544 | 39.0657 | 0.01 |
|  | Site | 0.2489 | 39.2268 | 0.01 |
| **GPS** | None | 0.2694 | 89.1020 | 0.01 |
|  | Age | 0.2722 | 88.2797 | 0.01 |
|  | Motion | 0.2604 | 89.1910 | 0.01 |
|  | Sex | 0.2125 | 94.1273 | 0.01 |
|  | Site | 0.2097 | 93.3001 | 0.01 |
| **Total** | None | 0.3093 | 338.9504 | 0.01 |
|  | Age | 0.3163 | 338.5785 | 0.01 |
|  | Motion | 0.3187 | 336.2625 | 0.01 |
|  | Sex | 0.2841 | 347.0704 | 0.01 |
|  | Site | 0.2787 | 348.6954 | 0.01 |

**Table S8:** *PANSS Score Prediction Performance for the median model in chronic psychosis***.** CPM results while controlling for key CP covariates for all PANSS summary measures. *Pearson’s correlation (r) and mean square error (MSE) between observed and predicted values

| **PANSS Score** | **Network** | **r*** | **MSE*** | **p-value** | **Z score** | **1-tailed p** |
| --- | --- | --- | --- | --- | --- | --- |
| **Positive** | Medial Frontal | 0.2682 | 26.92 | 0.02 | 0.733 | 0.232 |
|  | Frontoparietal | 0.2117 | 28.28 | 0.73 | 2.188 | 0.0143 |
|  | Default Mode | 0.3129 | 25.78 | 0.01 | -0.702 | 0.241 |
|  | Motor | 0.2063 | 28.29 | 0.1 | 1.524 | 0.064 |
|  | Visual I | 0.3252 | 25.36 | 0.01 | -1.146 | 0.126 |
|  | Visual II | 0.2973 | 26.17 | 0.01 | -0.173 | 0.431 |
|  | Visual Association | 0.301 | 25.85 | 0.03 | -0.294 | 0.384 |
|  | Limbic | 0.3214 | 25.35 | 0.01 | -1.113 | 0.133 |
|  | Subcortical | 0.314 | 25.91 | 0.01 | -0.76 | 0.224 |
|  | Cerebellum | 0.281 | 26.44 | 0.01 | 0.366 | 0.35732 |
| **Negative** |  | | | | | |
|  | Medial Frontal | 0.3105 | 38.13 | 0.01 | -0.545 | 0.293 |
|  | Frontoparietal | 0.2126 | 41.09 | 0.03 | 2.059 | 0.0198 |
|  | Default Mode | 0.3101 | 37.55 | 0.01 | -0.613 | 0.2704 |
|  | Motor | 0.2788 | 39.06 | 0.01 | 0.322 | 0.374 |
|  | Visual I | 0.3028 | 37.83 | 0.01 | -0.328 | 0.371 |
|  | Visual II | 0.2943 | 37.78 | 0.01 | -0.038 | 0.485 |
|  | Visual Association | 0.2852 | 38.31 | 0.01 | 0.262 | 0.397 |
|  | Limbic | 0.3097 | 37.67 | 0.01 | -0.64 | 0.261 |
|  | Subcortical | 0.3108 | 37.54 | 0.01 | -0.555 | 0.290 |
|  | Cerebellum | 0.289 | 38.08 | 0.01 | 0.134 | 0.447 |
| **GPS** | Medial Frontal | 0.2457 | 92.5292 | 0.01 | 0.82 | 0.206166 |
|  | Frontoparietal | 0.2499 | 91.8777 | 0.01 | 0.641 | 0.260646 |
|  | Default Mode | 0.2806 | 88.8048 | 0.01 | -0.428 | 0.33418 |
|  | Motor | 0.2959 | 87.2166 | 0.01 | -0.685 | 0.24652 |
|  | Visual I | 0.2619 | 90.0764 | 0.01 | 0.248 | 0.402145 |
|  | Visual II | 0.2673 | 89.6334 | 0.01 | 0.079 | 0.468612 |
|  | Visual Association | 0.2568 | 90.7608 | 0.01 | 0.406 | 0.342269 |
|  | Limbic | 0.2895 | 88.3784 | 0.01 | -0.713 | 0.237905 |
|  | Subcortical | 0.2609 | 90.7865 | 0.01 | 0.283 | 0.388589 |
|  | Cerebellum | 0.2616 | 89.9225 | 0.01 | 0.264 | 0.395855 |
| **Total** | Medial Frontal | 0.3077 | 345.50 | 0.01 | 0.06 | 0.476 |
|  | Frontoparietal | 0.2611 | 356.67 | 0.04 | 1.359 | 0.0871 |
|  | Default Mode | 0.3376 | 330.30 | 0.01 | -1.032 | 0.151 |
|  | Motor | 0.3005 | 341.61 | 0.01 | 0.196 | 0.422 |
|  | Visual I | 0.321 | 339.26 | 0.01 | -0.401 | 0.344 |
|  | Visual II | 0.3175 | 338.88 | 0.01 | -0.289 | 0.386 |
|  | Visual Association | 0.307 | 338.88 | 0.01 | 0.087 | 0.465 |
|  | Limbic | 0.326 | 340.75 | 0.01 | -0.559 | 0.288 |
|  | Subcortical | 0.3228 | 335.16 | 0.01 | -0.505 | 0.307 |
|  | Cerebellum | 0.3019 | 342.89 | 0.01 | 0.25 | 0.401 |

### **Table S9:** *Virtual lesion prediction performance for median model in chronic psychosis disorder*.

| **Dataset** | **Model** | **Data Applied** | **Pearson’s r** | **p value** | **T statistic** |
| --- | --- | --- | --- | --- | --- |
| HCP-EP | Total | Total CP | 0.0463 | 0.6112 | 0.5097 |
|  | Positive | Positive CP | 0.2292 | 0.0108 | 2.5905 |
|  | Negative | Negative CP | 0.0748 | 0.4110 | 0.8249 |
| SRPBS | Total | Total EP | 0.0707 | 0.4694 | 0.7261 |
|  | Positive | Positive EP | -0.0026 | 0.9789 | -0.0265 |
|  | Negative | Negative EP | -0.0001 | 0.9995 | -6.4632e-4 |
|  | General Psychopathology | General Psychopathology EP | 0.0531 | 0.5871 | 0.5447 |

**Table S10:** *Cross-Dataset External Validation*. Cross-dataset generalization experiments. HCP-EP = Human Connectome Project Early Psychosis, SRPBS = Strategic Research Program for Brain Sciences. EP = early psychosis, CP = chronic psychosis.

| **Dataset** | **Model** | **Data Applied** | **Pearson’s r** | **p value** | **T Statistic** |
| --- | --- | --- | --- | --- | --- |
| HCP-EP | Positive | Total CP | 0.2485 | 0.0056 | 2.82 |
|  |  | Negative CP | 0.2062 | 0.0221 | 2.32 |
|  |  | GPS CP | 0.2324 | 0.0097 | 2.63 |
|  | Negative | Positive CP | -0.0902 | 0.3211 | -0.93 |
|  |  | Total CP | -0.0165 | 0.8563 | -0.18 |
|  |  | GPS CP | -0.0327 | 0.7196 | -0.36 |
|  | Total | Positive CP | 0.0262 | 0.7736 | 0.29 |
|  |  | Negative CP | 0.1026 | 0.2589 | 1.13 |
|  |  | GPS CP | 0.0101 | 0.9116 | 0.11 |
| SRPBS | Positive | Negative EP | -0.0085 | 0.9306 | -0.087 |
|  |  | Total EP | 0.0165 | 0.8658 | 0.17 |
|  |  | GPS EP | 0.0444 | 0.6495 | 0.46 |
|  | Negative | Positive EP | 0.1938 | 0.0455 | 2.02 |
|  |  | Total EP | 0.1040 | 0.2865 | 1.07 |
|  |  | GPS EP | 0.0745 | 0.4455 | 0.77 |
|  | General Psych | Positive EP | -0.0232 | 0.8121 | -0.24 |
|  |  | Negative EP | 0.1128 | 0.2474 | 1.16 |
|  |  | Total EP | 0.0745 | 0.4455 | 0.77 |
|  | Total | Positive EP | -0.0378 | 0.6992 | -0.39 |
|  |  | Negative EP | 0.1495 | 0.1243 | 1.55 |
|  |  | GPS EP | 0.0178 | 0.8854 | 0.18 |

### **Table S11:** *Exploratory Cross-Symptom and Cross-Dataset External Validation*. The EP positive symptom predictive matrices had a positive association with CP total (r=0.2485, p=0.0056), negative (r=0.2062, p=0.0221), and general psychopathology scores (r=0.2324, p=0.0097). HCP-EP = Human Connectome Project Early Psychosis, SRPBS = Strategic Research Program for Brain Sciences.

| **HCP-EP (Edge Sign)** | **SRPBS (Edge Sign)** | **Overlap Count** | **Hygecdf p** |
| --- | --- | --- | --- |
| Negative (Positive) | Negative (Positive) | 6 | 0.074 |
| Negative (Negative) | Negative (Negative) | 2 | 0.60 |
| Positive (Positive) | Positive  (Positive) | 7 | 0.004 |
| Positive (Negative) | Positive (Negative) | 11 | 0.0016 |
| Total (Positive) | Total  (Positive) | 6 | 0.05 |
| Total (Negative) | Total  (Negative) | 4 | 0.24 |

**Table S12:** *Cross-Dataset Edge Comparison*. Edge-level comparisons, as measured by the hypergeometric cumulative (hygecdf) distribution function in Matlab. *p values are in parentheses. GPS = General Psychopathology, Neg = Negative, Pos = Positive. HCP-EP = Human Connectome Project Early Psychosis, SRPBS = Strategic Research Program for Brain Sciences.

| **HCP-EP (Edge Sign)** | **SRPBS (Edge Sign)** | **Correlation*** |
| --- | --- | --- |
| Negative (Positive) | Negative (Positive) | 0.2086 (0.0006) |
| Negative (Negative) | Negative (Negative) | 0.1928 (0.0015) |
| Positive (Positive) | Positive (Positive) | 0.0765 (0.2120) |
| Positive (Negative) | Positive (Negative) | 0.1733 (0.0044) |
| Total (Positive) | Total (Positive) | 0.4269 (0.0000) |
| Total (Negative) | Total (Negative) | 0.1341 (0.0281) |

**Table S13:** *Cross-Dataset Node Comparisons***.** Node-level comparisons, as measured by Pearson’s r across EP and CP for each symptom. HCP-EP = Human Connectome Project Early Psychosis, SRPBS = Strategic Research Program for Brain Sciences.

| **HCPEP (Edge Sign)** | **SRPBS (Edge Sign)** | **Pearson’s r (p value)** |
| --- | --- | --- |
| Negative (Positive) | Negative (Positive) | 0.7649 (0) |
| Negative (Negative) | Negative (Negative) | 0.5829 (0) |
| Positive (Positive) | Positive (Positive) | 0.6567 (0) |
| Positive (Negative) | Positive (Negative) | 0.7344 (0) |
| Total (Positive) | Total (Positive) | 0.6276 (0) |
| Total (Negative) | Total (Negative) | 0.6834 (0) |

**Table S14:** *Cross-Dataset Network Comparison***.** Network-level comparisons, as measured by Pearson’s r across EP and CP for each symptom. HCP-EP = Human Connectome Project Early Psychosis, SRPBS = Strategic Research Program for Brain Sciences.

| **Dataset** | **T-Test** | **Num components** | | **Statistic (corrected p value)** |
| --- | --- | --- | --- | --- |
| HCP-EP | Left | 12 | | 1 |
|  | Right | 11 | | 1 |
|  | Both | 8 | | 1 |
|  | | | Num Edges |  |
| SRPBS | Left | 1 | 9554 | 0 |
|  | Right | 1 | 11198 | 0 |

**Table S15:** *NBS – Case-Control (Control vs Patient)***.** n permutations = 5000, Z threshold = 1.98. Whole-brain cluster-based inference (NBS) on control versus patient connectomes for EP and CP. HCP-EP = Human Connectome Project Early Psychosis, SRPBS = Strategic Research Program for Brain Sciences.

| **PANSS Score** | **Feature Sign** | **Network Overlap*** | |
| --- | --- | --- | --- |
|  |  | **L** | **R** |
| **Total** | **Positive** | r=0.6872  (p=0) | r=0.5140  (p=0) |
|  | **Negative** | r=0.4753  (p=0) | r=0.6431  (p=0) |
| **Positive** | **Positive** | r=0.6463  (p=0) | r=0.4970  (p=0.0001) |
|  | **Negative** | r=0.5413  (p=0) | r=0.7061  (p=0) |
| **Negative** | **Positive** | r=0.6367  (p=0) | r=0.6559  (p=0) |
|  | **Negative** | r=0.5510  (p=0) | r=0.7401  (p=0) |

**Table S16:** *CP Case-Control NBS Network Overlap with EP Symptom Networks***.** NBS whole-brain analyses for EP symptoms and CP patient vs control differences. Overlap and similarity comparisons at the network levels are reflected as Pearson’s correlation and p values. *L=left, R=right. EP = early psychosis, CP = chronic psychosis.

| **PANSS Score** | **Feature Sign** | **Edge Overlap*** | | **Node Overlap**** | | **Network Overlap*** | |
| --- | --- | --- | --- | --- | --- | --- | --- |
|  |  | **L** | **R** | **L** | **R** | **L** | **R** |
| **Total** | **Positive** | r=0.0053 (p=0.1567) | r=-0.0068  (p=0.0673) | r=0.5144 (p=0) | r=0.4893  (p=0) | r=0.5933  (p=0) | r=0.5763  (p=0) |
|  | **Negative** | r=0.0063  (p=0.0894) | r=0.0029  (p=0.4358) | r=0.2870  (p=0) | r=0.2985  (p=0) | r=0.6674  (p=0) | r=0.6365  (p=0) |
| **Positive** | **Positive** | r=-0.0001 (p=0.9702) | r=0.0010  (p=0.7901) | r=0.3801 (p=0) | r=0.4055 (p=0) | r=0.6184  (p=0) | r=0.4775  (p=0.0002) |
|  | **Negative** | r=0.0008 (p=0.8317) | r=0.0131  (p=0.0004) | r=0.2972 (p=0) | r=0.2714 (p=0) | r=0.5354  (p=0) | r=0.7701  (p=0) |
| **Negative** | **Positive** | r=0.0004 (p=0.9202) | r=-0.0050  (p=0.1825) | r=0.4578 (p=0) | r=0.4247 (p=0) | r=0.5648  (p=0) | r=0.6463  (p=0) |
|  | **Negative** | r=-0.0044 (p=0.2359) | r=0.0084  (p=0.0240) | r=0.2205 (p=0.0003) | r=0.2790 (p=0) | r=0.4067  (p=0.0021) | r=0.6084  (p=0) |
| **General Psychopathology** | **Positive** | r=0.007 (p=0.8501) | r=-0.0049, (p=0.1924) | r=0.4473  (p=0) | r=0.4686  (p=0) | r=0.5818  (p=0) | r=0.6219  (p=0) |
|  | **Negative** | r=0.0011 (p=0.7623) | r=0.0058 (p=0.1178) | r=0.2281  (p=0.0002) | r=0.2635 (p=0) | r=0.5635  (p=0) | r=0.6948  (p=0) |

### **Table S17:** *CP Case-Control NBS Network Overlap with CP Symptom Networks***.** NBS whole-brain analyses for CP symptoms and patient vs control differences. Overlap and similarity comparisons at the edge, node, and network levels are reflected as Pearson’s correlation and p values. L = left-tailed p-test (controls < patients), R = right-tailed p-test (patients < controls), *p values are in parentheses, **fractional overlap, then p value. CP = chronic psychosis.

| **Canonical Network** | **Nodes** | **Total Nodes in Network** | **% missing in Network** |
| --- | --- | --- | --- |
| 2 | 112, 116, 242, 246 | 34 | 11.76 |
| 3 | 115, 239 | 20 | 10 |
| 4 | 109 | 50 | 2 |
| 6 | 100 | 9 | 11.11 |
| 7 | 240 | 18 | 5.56 |
| 10 | 104, 105, 107, 108, 118, 129, 131, 236, 237, 243, 249, 250, 252, 256, 266, 267, 268 | 31 | 54.84 |

**Table S18:** *Exclusion of Missing Nodes from CP Sample.* 37 CP patients had missing nodes (~30%). 26 nodes total across these 37 subjects were missing (had no functional value). These nodes were removed from all participants to keep the same amount of node coverage consistent across participants. EP = early psychosis, CP = chronic psychosis.
